# Economic Losses Associated with COVID-19 Deaths in the United States

**DOI:** 10.1101/2020.10.25.20219212

**Authors:** Troy Quast, Ross Andel, Sean Gregory, Eric A. Storch

**Affiliations:** University of South Florida, College of Public Health, Tampa, FL 33612; University of South Florida, College of Behavioral and Community Sciences, Tampa, FL 33620; Charles University and Motol University Hospital, Department of Neurology, Prague; International Clinical Research Center, St. Anne’s University Hospital, Brno; Department of Politics & International Affairs, Northern Arizona University, Flagstaff, AZ 86011; Menninger Department of Psychiatry & Behavioral Sciences, Baylor College of Medicine, Houston, TX 77030

**Keywords:** COVID-19, economic loss, years of life lost, United States

## Abstract

In addition to the overwhelming health effects of COVID-19, the disease has inflicted unprecedented economic damage. Vast resources have been directed at COVID-19 testing and health care while economic activity has been substantially curtailed due to disruptions resulting from individual choices and government policies. This study estimates the economic loss associated with COVID-19 deaths in the U.S. from February 1, 2020 through July 11, 2020. We use estimates of years of life lost that are based on the age and gender of decedents. Using a value of life year estimate of $66,759, we calculate economic losses of roughly $66 billion. The losses are concentrated in New York and New Jersey, which account for 17.5% of the total losses. Our analysis of per capita losses by state indicates that the highest values are located in the northeastern region of the country, while the values in the western states are relatively low. While economic losses associate with COVID-19 deaths is just one aspect of the pandemic, our estimates can provide context to the value of prevention and mitigation efforts.

**JEL codes:** I12, I18, J17

## Introduction

While the health effects of COVID-19 are devastating, the economic effects of the pandemic are also profound. Estimates suggest that if 20% of the population were infected, the direct medical costs in the United States (U.S.) would be roughly $163 billion (Bartsch et al, 2020). These costs include testing, ambulatory care, hospitalizations, and extended treatment of long-term effects of the disease. The economic costs also include the reduction and, in some cases, complete stoppage of economic activity due to government policies and individual behavior. These costs have been estimated to reduce real gross domestic product (GDP) growth by 5% per month (Makridis and Hartley, 2020).

Another important component of the economic costs of COVID-19 involves losses due to deaths from the disease. While the approaches are imperfect, economists have long attempted to estimate costs of deaths due to actual or potential policies and events. A common approach is employ value of a statistical life (VSL) estimates. Generally, the number of deaths is multiplied by the VSL to estimate the economic cost. This approach is used by U.S. government agencies (Robinson, 2007) and by researchers in areas such as transportation (Rizzi and Ortuzar, 2005; Dua, 2015), health interventions (Laxminarayan et al, 2014; Sproul et al, 2012; Scharff, 2012), and environmental studies (Franchini, 2015; Lee et al, 2011). While these analyses are informative, a significant drawback is that they do not explicitly account for the age profile of the decedents. For instance, under this approach the same value is assigned to a newborn as is assigned to a very sick individual near death.

This paper estimates the economic losses associated with the roughly 130,000 COVID-19 deaths in the U.S. from February 1, 2020 to July 11, 2020. Our approach employs years of lost life (YLL) estimates, which are based on the age and gender of the decedents, reported in Quast, et al (2020). We estimate the economic losses associated with COVID-19 deaths to be roughly $66 billion.

## Methods and Materials

Our general approach is to multiply the YLLs by an estimated value of each life year. This strategy has been used in studies of areas including health behaviors (Nonnemaker et al, 2014), pollution (Desaigues et al, 2011; Grandjean and Bellanger, 2017; Kim et al, 2020), and health outcomes (Temkin et al, 2019; Pezzullo, et al, 2018). Our YLL estimates are based on an analysis by Quast et al (2020) of data contained from the “Provisional COVID-19 Death Counts by Sex, Age, and State” published by the National Center for Health Statistics in the U.S. Centers for Disease Control and Prevention (CDC). These data separately report deaths for New York and New York City and do not contain data for Wyoming. Quast et al (2020) merged the death counts with life expectancy estimates by age and gender obtained from actuarial life tables published by the U.S. Social Security Administration. For each age-gender group, the unadjusted YLL estimates were obtained by multiplying the number of individuals in that group by the difference in life expectancy and the age at death. The population-wide number of YLLs was found by summing across the age-gender groups.

As noted by Hanlon et al (2020), those who died of COVID-19 often had pre-existing morbidities which may have otherwise prevented them from reaching their life expectancy. The authors created a model of COVID-19 deaths in the United Kingdom that used data from Italy and found that, adjusting for pre-existing morbidities, the average estimated YLLs for men fell from 14 to 13 and for women from 12 to 11. Our primary specification follows Quast et al (2020) and reduces the life expectancy of decedents by 25% to reflect the typically lower pre-COVID-19 health status of those who died.

A value of life year (VOLY) estimate must be identified to calculate the economic cost associated with YLLs. We employ a value reported by Desaigues, et al (2011) that was based on a stated preferences approach. They asked residents of European Union countries their willingness to pay for policies that reduced pollution and, thus, increased life expectancy. Their estimate of 41,000 Euros has been employed in previous economic loss calculations (Temkin et al, 2019; Grandjean and Bellanger, 2017; Kim et al, 2020). The value converted to 2020 U.S. dollars is $66,759. We also calculated economic losses using lower and upper benchmarks ($100,000 and $150,000) employed by the Institute of Clinical and Economic Review when conducting cost-effectiveness analyses (Institute for Clinical and Economic Review, 2017). However, these benchmark values are based on quality-adjusted life years and are not directly comparable to VOLY estimates. We discounted future dollar amounts by 3% per year.

To estimate per-capita values, we use population data from the “Annual State Resident Population Estimates” published by the U.S. Census Bureau as of July 1, 2019 and were obtained by gender. We obtained the population of New York City and the percentage of population by gender as of July 1, 2019 from “QuickFacts: New York City, New York” published by the U.S. Census Bureau.

## Results

Table 1 details the economic losses using our primary VOLY estimate of $66,759 and the two benchmark ICER values. The values are based on reducing the life expectancy estimates by 25%. Appendix Table 1 reports the values for our primary VOLY estimate and life expectancy reductions of 0% and 50%.

**Table 1.** Economic losses ($000s) by jurisdiction based on various VOLY estimates.

| **Jurisdiction** | **VOLY estimate** |  |  |
| --- | --- | --- | --- |
| | **\$66,759** | **\$100,000** | **\$150,000** |
| US Total | 65,824,441 | 98,600,100 | 147,900,150 |
| New York City | 11,600,711 | 17,377,000 | 26,065,500 |
| New Jersey | 6,971,909 | 10,443,400 | 15,665,100 |
| New York (excl NYC) | 5,467,362 | 8,189,700 | 12,284,550 |
| California | 3,790,242 | 5,677,500 | 8,516,250 |
| Illinois | 3,506,650 | 5,252,700 | 7,879,050 |
| Pennsylvania | 3,159,436 | 4,732,600 | 7,098,900 |
| Massachusetts | 3,106,296 | 4,653,000 | 6,979,500 |
| Michigan | 2,921,841 | 4,376,700 | 6,565,050 |
| Texas | 2,206,318 | 3,304,900 | 4,957,350 |
| Florida | 2,128,410 | 3,188,200 | 4,782,300 |
| Maryland | 1,848,957 | 2,769,600 | 4,154,400 |
| Louisiana | 1,664,569 | 2,493,400 | 3,740,100 |
| Connecticut | 1,692,207 | 2,534,800 | 3,802,200 |
| Arizona | 1,376,370 | 2,061,700 | 3,092,550 |
| Georgia | 1,381,978 | 2,070,100 | 3,105,150 |
| Indiana | 1,268,421 | 1,900,000 | 2,850,000 |
| Ohio | 1,237,912 | 1,854,300 | 2,781,450 |
| Virginia | 983,160 | 1,472,700 | 2,209,050 |
| Colorado | 766,660 | 1,148,400 | 1,722,600 |
| Alabama | 707,378 | 1,059,600 | 1,589,400 |
| Mississippi | 691,156 | 1,035,300 | 1,552,950 |
| North Carolina | 615,919 | 922,600 | 1,383,900 |
| Minnesota | 615,651 | 922,200 | 1,383,300 |
| Washington | 580,737 | 869,900 | 1,304,850 |
| South Carolina | 514,712 | 771,000 | 1,156,500 |
| Missouri | 467,046 | 699,600 | 1,049,400 |
| Wisconsin | 424,788 | 636,300 | 954,450 |
| District of Columbia | 389,071 | 582,800 | 874,200 |
| Iowa | 395,547 | 592,500 | 888,750 |
| Tennessee | 375,586 | 562,600 | 843,900 |
| Rhode Island | 383,530 | 574,500 | 861,750 |
| Nevada | 331,859 | 497,100 | 745,650 |
| New Mexico | 300,816 | 450,600 | 675,900 |
| Kentucky | 311,097 | 466,000 | 699,000 |
| Delaware | 245,940 | 368,400 | 552,600 |
| Oklahoma | 208,622 | 312,500 | 468,750 |
| Arkansas | 198,675 | 297,600 | 446,400 |
| Nebraska | 156,884 | 235,000 | 352,500 |
| Kansas | 157,551 | 236,000 | 354,000 |
| New Hampshire | 145,601 | 218,100 | 327,150 |
| Utah | 124,706 | 186,800 | 280,200 |
| Oregon | 124,038 | 185,800 | 278,700 |
| South Dakota | 55,877 | 83,700 | 125,550 |
| Maine | 53,073 | 79,500 | 119,250 |
| North Dakota | 45,797 | 68,600 | 102,900 |
| Idaho | 46,331 | 69,400 | 104,100 |
| West Virginia | 43,994 | 65,900 | 98,850 |
| Vermont | 22,231 | 33,300 | 49,950 |
| Montana | 10,815 | 16,200 | 24,300 |

The total estimated losses associated with COVID-19 deaths in the U.S. during our analysis period were roughly $66 billion. The losses were highly concentrated among the top states. New York City alone accounted for roughly 17.5% of the total losses while New York City, New Jersey, and New York state (excluding New York City) combined incurred over one-third of the total losses. The top ten jurisdictions accounted for roughly 70% of the total losses.

Figure 1 shows the economic loss per capita using our primary VOLY estimate and the 25% reduction in estimated life expectancy. (New York and New York City are combined in the map.) The largest values are concentrated in the northeastern section of the country, while the highest value outside of the region was in Louisiana. The western half of the country had lower per capita economic losses.

**Figure 1.**
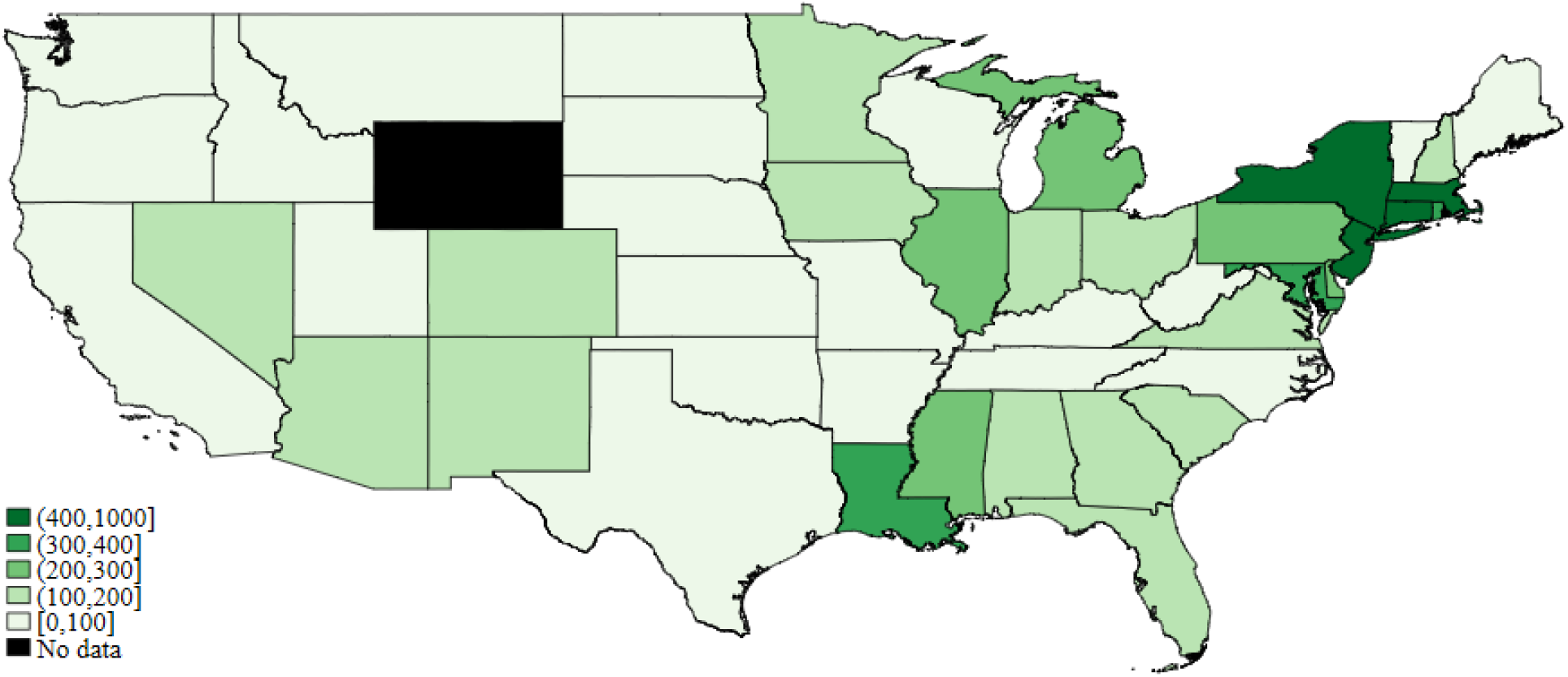
Economic loss ($) per capita by jurisdiction.

## Discussion

The human toll of COVID-19 is devastating and takes precedence when considering the pandemic’s effects. However, estimates of the economic loss can provide context in terms of the cost of prevention and mitigation efforts. Deaths are just one aspect of the toll of COVID-19, but are arguably the most important. We estimate the total economic loss due associated with COVID-19 deaths in the U.S. to be roughly $66 billion which, if extrapolated to an annual basis, corresponds to roughly 0.7% of the U.S. GDP in 2019. We observe large absolute and per capita losses in the northeast region of the country while the western region of the country generally has lower per-capita losses.

As noted above, another potential approach to estimate economic losses due to deaths is to base the calculation on the VSL rather than on the projected number of years of life lost. VSLs are typically estimated via either stated or revealed preference for how much compensation an individual would demand to incur greater risk. Using a VSL of $9.6 million (Viscusi, 2015; Viscusi and Masterman, 2017), the corresponding economic loss associated with the 130,088 deaths is roughly $1.2 trillion, a value roughly eighteen times greater than our estimate. Economic loss estimates based on VOLYs and VSLs are not directly comparable, but the wide range of the two values suggest that adjusting for the relatively older age profile of COVID-19 decedents has a substantial effect on economic loss estimates.

Our analysis has several limitations. Most notably, we only estimate losses associated with deaths. Costs due to other aspects such as hospitalizations, long-term health effects, and reductions in economic activity need to be included in an estimate of the overall cost of COVID-19. Our reduction in projected life expectancy by 25% is an approximate value. However, we believe it is a conservative approach that reflects the current understanding of COVID-19 mortality risk. We used a single VOLY estimate for the entire country which does not reflect potential variation in valuations across states. Further, if an estimate was available, it would be preferred to use an estimate based on surveys of U.S. rather than E.U. residents. Finally, our analysis period only represents the early stages of the COVID-19 outbreak in the

U.S. As the virus ebbs and flows throughout the country and additional lives are lost, both the total economic loss but also the ranking of jurisdictions will change.

## Compliance with Ethical Standards

The authors declare that they have no conflict of interest.

## Data Availability

The study data were obtained from the U.S. Centers for Disease Control's report, "Provisional COVID-19 Death Counts by Sex, Age, and State" (https://data.cdc.gov/NCHS/Provisional-COVID-19-Death-Counts-by-Sex-Age-and-S/9bhg-hcku).

---

**Appendix Table 1.** Economic losses ($000s) by jurisdiction based on various adjustments to life expectancy.

| Jurisdiction | Reduction in life expectancy |  |  |
| --- | --- | --- | --- |
|  | 0% | 25% | 50% |
| US Total | 82,794,044 | 65,824,441 | 46,658,933 |
| New York City | 14,521,418 | 11,600,711 | 8,264,564 |
| New Jersey | 8,766,191 | 6,971,909 | 4,943,905 |
| New York (excl NYC) | 6,891,131 | 5,467,362 | 3,866,748 |
| California | 4,748,701 | 3,790,242 | 2,698,265 |
| Illinois | 4,398,751 | 3,506,650 | 2,493,048 |
| Pennsylvania | 4,005,540 | 3,159,436 | 2,220,404 |
| Massachusetts | 3,956,539 | 3,106,296 | 2,172,271 |
| Michigan | 3,672,546 | 2,921,841 | 2,072,466 |
| Texas | 2,750,404 | 2,206,318 | 1,579,117 |
| Florida | 2,680,174 | 2,128,410 | 1,507,084 |
| Maryland | 2,322,546 | 1,848,957 | 1,312,615 |
| Connecticut | 2,150,841 | 1,692,207 | 1,185,974 |
| Louisiana | 2,088,088 | 1,664,569 | 1,183,370 |
| Georgia | 1,733,064 | 1,381,978 | 982,692 |
| Arizona | 1,718,310 | 1,376,370 | 983,827 |
| Indiana | 1,605,287 | 1,268,421 | 893,035 |
| Ohio | 1,567,835 | 1,237,912 | 870,804 |
| Virginia | 1,241,584 | 983,160 | 693,826 |
| Colorado | 968,339 | 766,660 | 541,015 |
| Alabama | 886,359 | 707,378 | 503,430 |
| Mississippi | 863,928 | 691,156 | 493,215 |
| Minnesota | 782,883 | 615,651 | 431,263 |
| North Carolina | 774,872 | 615,919 | 436,537 |
| Washington | 733,281 | 580,737 | 409,900 |
| South Carolina | 647,562 | 514,712 | 364,571 |
| Missouri | 590,951 | 467,046 | 328,922 |
| Wisconsin | 533,805 | 424,788 | 301,417 |
| Iowa | 498,022 | 395,547 | 280,054 |
| Rhode Island | 488,476 | 383,530 | 268,171 |
| District of Columbia | 485,471 | 389,071 | 278,118 |
| Tennessee | 470,584 | 375,586 | 267,303 |
| Nevada | 415,508 | 331,859 | 236,327 |
| Kentucky | 393,010 | 311,097 | 219,504 |
| New Mexico | 373,383 | 300,816 | 216,499 |
| Delaware | 310,162 | 245,940 | 173,840 |
| Oklahoma | 263,297 | 208,622 | 147,270 |
| Arkansas | 248,210 | 198,675 | 141,863 |
| Kansas | 198,074 | 157,551 | 111,688 |
| Nebraska | 196,672 | 156,884 | 111,554 |
| New Hampshire | 186,057 | 145,601 | 101,407 |
| Oregon | 156,550 | 124,038 | 87,521 |
| Utah | 156,216 | 124,706 | 88,789 |
| South Dakota | 70,498 | 55,877 | 39,455 |
| Maine | 67,560 | 53,073 | 37,118 |
| Idaho | 59,282 | 46,331 | 32,245 |
| North Dakota | 57,947 | 45,797 | 32,178 |
| West Virginia | 56,011 | 43,994 | 30,709 |
| Vermont | 28,373 | 22,231 | 15,488 |
| Montana | 13,752 | 10,815 | 7,544 |

